# Ascertainment rate of SARS-CoV-2 infections from healthcare and community testing in the UK

**DOI:** 10.1101/2021.02.09.21251411

**Authors:** Ewan Colman, Gavrila A. Puspitarani, Jessica Enright, Rowland R. Kao

## Abstract

The proportion of SARS-CoV-2 infections ascertained through healthcare and community testing is generally unknown and expected to vary depending on natural factors and changes in test-seeking behaviour. Here we use population surveillance data and reported daily case numbers in the United Kingdom to estimate the rate of case ascertainment. We mathematically describe the relationship between the ascertainment rate, the daily number of reported cases, population prevalence, and the sensitivity of PCR and Lateral Flow tests as a function time since exposure. Applying this model to the data, we estimate that 20-40% of SARS-CoV-2 infections in the UK were ascertained with a positive test with results varying by time and region. Cases of the Alpha variant were ascertained at a higher rate than the wild type variants circulating in the early pandemic, and higher again for the Delta variant and Omicron BA.1 sub-lineage, but lower for the BA.2 sub-lineage. Case ascertainment was higher in adults than in children. We further estimate the daily number of infections and compare this to mortality data to estimate that the infection fatality rate increased by a factor of 3 during the period dominated by the Alpha variant, and declined in line with the distribution of vaccines.

---

Testing for SARS-CoV-2 in the UK aims to accomplish two things - first, to rapidly confirm suspected cases of COVID-19 disease via symptomatic testing in order to contain outbreak clusters, and second, to establish the overall burden of infection by taking a random sample of the population. Since not all infected individuals receive a test, and some of those who do will receive a false negative result, the number of positive diagnostic tests provides a *lower* estimate of the number of people exposed to the virus [1]. In contrast, random testing can provide an unbiased estimate of prevalence, but is an inefficient way to rapidly identify infection clusters, and may also have biases depending on the extent to which a positive test indicates the true infection status of the individual.

Both types of data are available in the UK: the number of positive tests from people with suspected infection are published daily on the UK government dashboard [2], and the Office for National Statistics COVID-19 Infection Survey (CIS) regularly publishes estimates of the population prevalence based on unbiased sampling [3]. The existence of these sources creates an opportunity to answer an important question: *what proportion of all infections are being reported through diagnostic testing?* Knowing this can help to estimate true incidence rates, a quantity central to understanding how the virus is spreading, and determine the infection fatality rate (IFR) of the disease.

Here we describe how diagnostic case numbers can be used to model the proportion of the population testing positive. By calibrating this model against surveillance data, we estimate the case ascertainment rate, defined as the proportion of infections that were reported through diagnostic testing; the incidence, defined as the number of newly infected individuals each day; and the IFR. Our differs from previous work as we do not rely on prior assumptions about the IFR to estimate incidence [4, 5]. Moreover, we estimate the IFR using a data set substantially larger than any that has previously been used [6].

The model incorporates the different types of test and differences caused by variants of SARS-CoV-2, which have been shown to result in higher severity and a different range of clinical symptoms [7, 8]. New variants might have different pathological characteristics that could potentially affect the test-seeking behaviour of those infected, which we expect to directly affect case ascertainment. Examination of age related and regional variation in case ascertainment provides a novel way to consider these developments, and to enrich our understanding of the epidemiology of the virus.

## 1 Methods

### 1.1 Data

We are primarily concerned with daily Pillar 1 and 2 case data [2], hereafter referred to as diagnostic test cases, which represent tests done in health care settings and the community, respectively. We use *C*_*q*_(*t*) to denote the number of Pillar 1 and 2 cases from test type *q* on day *t*. Here, *q* can be PCR or LFD. These counts come from lab-based PCR tests and lateral flow device (LFD) testing, as performed in many community settings [9]. We use data provided for the 12 regions of the UK (9 regions of England and 3 other nations), and the 19 5-year age bands, which we aggregate into 7 distinct age bands to be consistent with the CIS data. Since the age bands for the the Pillar 1 & 2 data are not perfectly aligned with those for the CIS data, we first distribute them into 1-year age brackets, assuming an equal distribution of cases within each bracket, before re-aggregating.

At the time of writing, the number of cases detected by test type were available for England, but not provided at the regional level or for different age bands. We therefore approximate the proportion of cases that come from each test type by partitioning the total case numbers according to the the proportion calculated at the national scale.

The CIS provides estimates for the estimated percentage of people testing positive for coronavirus for the 9 regions of England, 3 other nations of the UK, and 7 age bands in England. Data for nations and age bands represent samples collected over 14-day intervals. We take the 7th day as the representative time point of this estimate. Data for the nations is provided weekly and so we take it to represent the 4th day of the 7-day period. Population counts for the 9 regions and the 7 age categories were compiled from CIS data [10].

There is uncertainty in the CIS which we transfer to our own analysis as follows. From the CIS, we use the “rate” and 95% confidence interval over a series of time points between September 2020 and June 2022 [3]. The exact distributions are not provided by the CIS source, so we approximate them with Normal distributions with mean equal to the CIS rate values and variance calculated to be as consistent as possible with the CIS confidence intervals. We construct a sampled time series by taking a series of samples from the series of distributions. The sampled time series of percentages is then applied to the population to give, *M*(*t*), the total number of test-positive people, where *t* is the midpoint of the time interval that the data represent. We repeat our analysis on 200 time series independently constructed in this way to obtain a distribution of results.

The CIS provides an estimate of the proportion of tests that achieve different testing targets using the TaqPath test [11]. We use these to estimate the proportion of infections that are from wild-type, Alpha, Delta and Omicron variants (BA.1 and BA.2 sub-lineages) of SARS-CoV-2. We consider tests that are negative for the S target gene and positive for the two other targets, known as S-gene target failure (SGTF), to be a proxy for the Alpha and Omicron BA.1 variants. Since tests that are positive for S and exactly one of the other targets (N or ORFab1) may indicate any lineage [12], we discard those that are negative on the S target and one other target from our calculation of the SGTF proportion.

Based on the time points when the SGTF proportion reached locally maximum or minimum values, we assume the variant class follows from SGTF as follows. All infections from the beginning of the pandemic until November 1st 2020, and all infections up to March 1st 2021 that are not SGTF, are of the wild-type variants. Infections that are SGTF are of the Alpha variant if they were reported between November 1st 2020 and November 1st 2021, and the Omicron BA.1 sub-lineage if they we reported after November 1st 2021. Infections that are not SGTF are assumed to be the Delta variant if they were reported between March 1st 2021 and and January 9th 2022, and the Omicron BA.2 sub-lineage if they we reported after January 9th 2022. We therefore consider 5 variant classes of interest: Wild-type, Alpha, Delta, Omicron BA.1 and omicron BA.2 whose proportions we denote using *p*_*w*_, *p*_*A*_, *p*_Δ_, *p*_*o*1_, *p*_*o*2_, respectively.

The number of deaths in England of individuals who have tested positive for coronavirus within 28 days is provided in 5-year age bands [13]. As with the case numbers, these data were first distributed into 1-year age brackets assuming an equal distribution of cases within each bracket, before being re-aggregated into age bands consistent with the CIS.

#### Modelling the time from exposure to the time of positive test

We define two random variables, *X* and *T*, representing the time an individual was exposed and the time they received a positive test, respectively. Assuming daily time steps, we express the probability that an individual who received a positive test from a sample taken at time *t*^+^ was first exposed to the virus at time *x*,

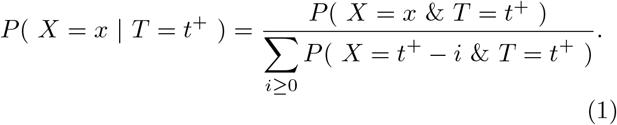

The joint probability distribution *P* (*X* = *x & T* = *t*^+^) can be pieced together from various sources by considering the sequence of events that result in an individual testing positive.

First, we consider the time the individual was exposed to the virus and acquired the infection. We denote the prior probability that the infection was acquired at time *x* by *B*(*x*). Next, we consider the time between exposure and the time that they received a test. For symptomatic cases we assume that the test occurs shortly after symptom onset, i.e. the time since exposure *τ* = *t*^+^ − *x* is equal to the sum of the incubation period and a delay parameter *δ*_*k*_ that we assume to be a fixed quantity; *k* ∈ {PCR, LFD} represents the type of test being performed. The probability of a test on day *t*^+^ is thus *R*(*t*^+^ − *δ*_*k*_ − *x*) where *R*(*i*) is the probability that the duration of the incubation period is *i*, which we assume to be Log-normal with a mean of 5.5 days and dispersion parameter 1.52 [15, 16]. To get a probability distribution expressing the length of the incubation period in discrete days, we integrate over consecutive intervals of length 1.

Once the individual has acquired the infection and has had a test, the test must be positive to become an ascertained infection. The probability of testing positive varies as a function, *S*_*k*_(*τ*), of the time since exposure *τ* = *t*^+^− *x*. We use the functions provided by Hellewell et al. [14] and shown in Fig. 1. The PCR curve is similar in shape to the shedding profile found in other studies [17–20] with viral load typically peaking at day 3-5 and persisting for a mean duration of 17 days [21]. Variation is associated with severity of illness but not age or sex [22–24]. Studies that look for a difference between asymptomatic and symptomatic infections do not report consistent results [18, 25]. While one study with a small sample found that the Alpha variant had a longer viral course than the wild type [26], studies generally show that shedding profiles do not differ significantly between variants [27,28]. In contrast, vaccination has been shown to reduce incidence of high shedding rates and duration of shedding [28–31] which we address in a later section.

**Figure 1:**
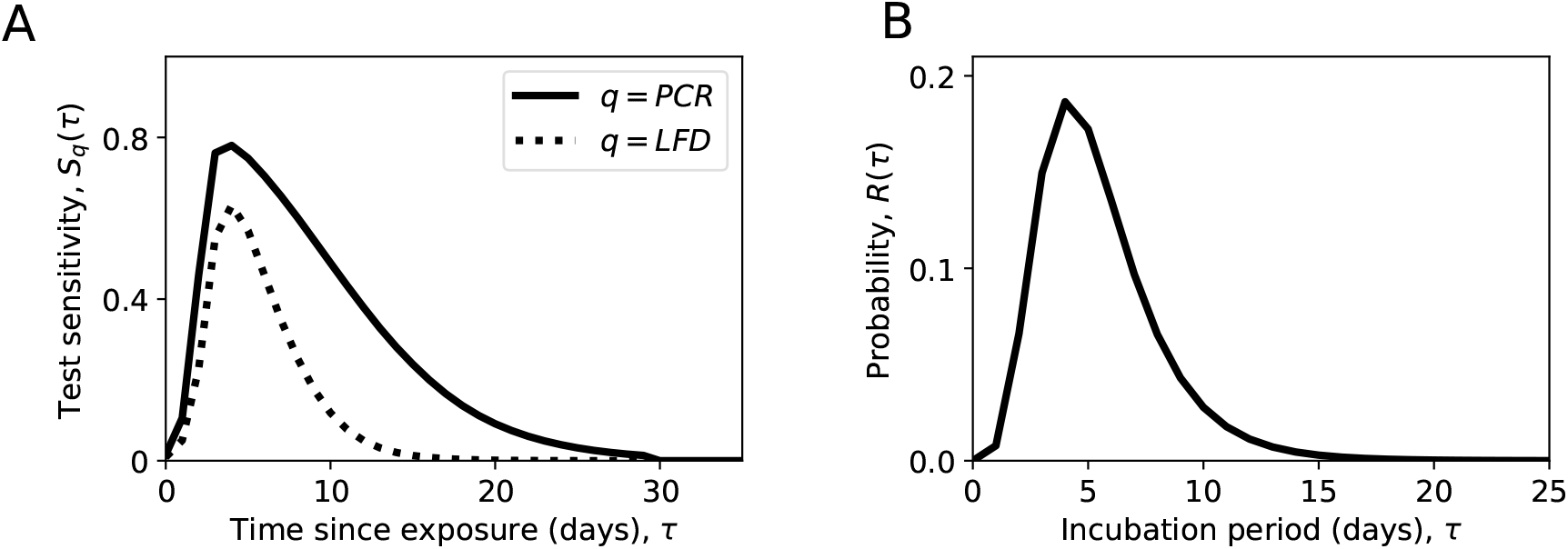
(A) The test sensitivity as a function of time from Hellewel et al. [14]. The function, *S*(*τ*) gives the probability that a PCR test will be positive if performed on an infected person *τ* days after exposure. (B) The incubation period probability distribution from Lauer et al. [15], shown here are the probability of symptom onset on each day since exposure.

We express the probability that an infected individual was exposed on day *x* and tested positive on day *t*^+^ by multiplying the probabilities mentioned above,

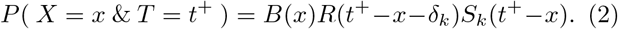

If we assume an uninformative prior probability, *B*(*x*), of exposure on day *x*, then substituting Eq. (2) into Eq. (1) gives

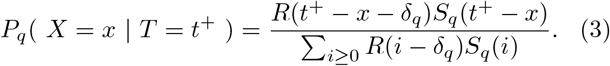

Substituting *τ* = *t*^+^ − *x*, and using the notation *P*_*q*_(*τ*) = *P*_*q*_(*X* = *t*^+^ − *τ* |*T* = *t*^+^), we have

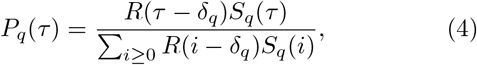

giving the probability distribution of time between exposure and test for the set of ascertained cases corresponding to the test types *q* ∈ {PCR, LFD}.

We justify our choice to disregard the form of *B*(*x*) by examining the the effect it has on *P*_*q*_(*τ*) in the most extreme feasible scenarios. Supposing *B*(*x*) = *Ag*^*x*^ where *A* is a constant and *g* is the daily multiplicative growth rate, Eq.(1) can be solved to get a revised version of *P*_*q*_(*τ*) that now also depends on *g* (*A* does not appear in the solution). The largest observed daily growth rate in the 7-day average of cases, *C*_*PCR*_(*t*) + *C*_*LFD*_(*t*), is *g* = 1.15. Under the revised formula, the mean values of *P*_*LFD*_(*τ*) and *P*_*LFD*_(*τ*) are respectively 0.58 and 0.35 days shorter than when Eq.(4) is used. The lowest growth rate observed is *g* = 0.85, which results in the mean values of *P*_*LFD*_(*τ*) and *P*_*LFD*_(*τ*) to be 0.98 and 0.52 days longer, respectively. In general the effect is considerably smaller than in these examples so we proceed under the assumption that Eq.(4) is sufficient for our analysis.

#### Estimating the ascertainment rate

We define the *ascertainment rate* as the proportion of SARS-CoV-2 infections that result in a positive PCR or LFD test and are recorded in the Pillar 1 & 2 case data. We introduce the time-dependent ascertainment rate ***θ***, a vector whose *x*th element, *θ*_*x*_, is the proportion of infections that occurred at time *x* that get reported through diagnostic testing at any subsequent time. We also consider the incidence, *I*(*x*), defined as the number of newly acquired infections on day *x*.

The number of ascertained cases that were exposed at time *x* can be expressed in two ways: first, by multiplying the incidence by the ascertainment rate, and second by expressing the number of reported cases that were exposed on day *x* as a function of the daily case counts. Equating the two gives

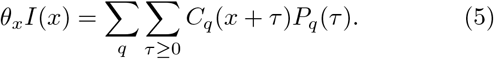

We estimate the number of individuals in the population who would test positive (by PCR) on day *t*, if tested, by summing over all infections times and weighting by the the probability that each one is test-positive on day *t*

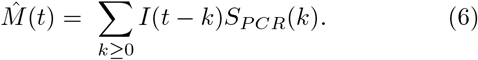

Combining with Eq. (5) we can express this as a function of time and the unknown vector of parameters ***θ***

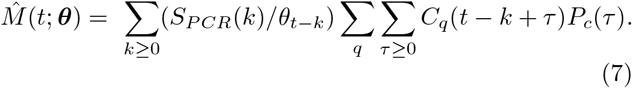

We estimate ascertainment by finding the vector ***θ*** that minimizes the difference between the estimated and observed values,

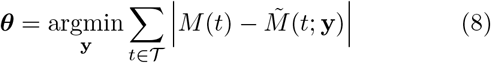

where 𝒯 is the set of time points for which we have empirical estimates of prevalence. Eq. 8 combines the daily diagnostic case counts, the population positivity from surveil-lance, the incubation period distribution, and the time-dependent test sensitivity of PCR and LFD tests, to provide an estimate of the proportion of infections being reported at time *t*.

In practice, we estimate *θ*_*x*_ at weekly time points and use linear interpolation to create a daily time series. The solution to Eq. (8) is found numerically. The optimisation is made more efficient by inputting an initial guess based on an approximation to *θ* given by

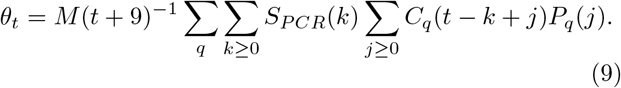

Note that this equation uses the reported prevalence shifted forward by 9 days which is approximately the time since exposure of someone who received a positive test result through random surveillance. We estimate the credible interval for *θ* by substituting the upper and lower bounds of the credible interval for *M*(*t*) into Eq (8).

#### Time-independent ascertainment rate

Motivated by the possibility that variants of concern may have different pathological characteristics to each other or elicit different test-seeking responses, we estimate ascertainment rates for each variant class. Unlike the previous section these rates are constant for each variant in each region and age band. We let 𝒱 = {*w, A*, Δ, *o*1, *o*2} to denote the wild-type, Alpha, Delta, Omicron BA.1 and Omicron BA.2 variants, respectively, and ***ϕ*** = (*ϕ*_*w*_, *ϕ*_*A*_, *ϕ*_Δ_, *ϕ*_*o*1_, *ϕ*_*o*2_) where *ϕ*_*v*_ denotes the time-independent rate for variant class *v*. Recalling that *p*_*v*_(*t*) is the proportion of infections caused by variants of class *v*, we use a revised estimate of *M*(*t*) that weights the contribution of each variant class by its proportion

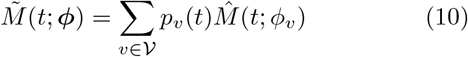

We can then estimate by taking the value that minimizes the absolute error between *M*(*t*) and 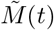 taking only the time points 𝒱^*′*^ up to March 1st 2021 when SGTF positive tests were associated with the Delta variant. Specifically, the time-independent ascertainment rates are estimated by numerically solving

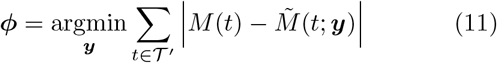

#### Infection fatality rate

The infection fatality rate (IFR) is defined as the proportion of individuals infected who then die as a direct result of the infection. For a given monthly mortality figure, we count the corresponding number of infections from *I*(*x*) summed over all corresponding exposure dates. We include a 21 day time from exposure to death (5 days to symptom onset and 16 days between onset and death) to be consistent with previous studies and the time between peaks in case and death data in England [32]. For example, the IFR for September is the number of recorded deaths in that month divided by the number of infections that occurred between August 10th and September 10th.

#### Effect of vaccination on ascertainment rate

We apply different test sensitivity functions to the proportion of infections that are in individuals who have received a vaccine and those who have not. Here we first describe how the proportion of infections that are in vaccinated, and unvaccinated, people is estimated. We then describe how this was accommodated into our analysis.

Vaccine effectiveness is defined as

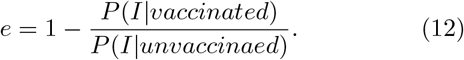

It varies depending on the time since vaccination, the number of doses, the specific vaccine given, and the outcome measured e.g. infection, symptoms, hospitalisation. Effectiveness is lowest when the measured outcome is infection of any kind, regardless of symptoms. This is estimated to be *e* = 0.56 (56%) [29]. We choose to use this low value to avoid underestimating the effect of vaccines; lower estimates of effectiveness results in higher proportions of infections that are subject to the effects of the vaccine.

We want to know the proportion of infections on day *t* that are in the population of people that have received the vaccine by day *t*, which we denote with *π*(*t*). We have that *P* (*I*|*vaccinated*) = *π*(*t*)*I*(*t*)*/V* (*t*) where *I*(*t*) is the number of new infections on day *t* and *V* (*t*) is the number of people vaccinated by day *t*. Similarly *P* (*I*|*unvaccinated*) = (1 − *π*(*t*))*I*(*t*)*/*(*N* − *V* (*t*)) where *N* is the population. Substituting into Eq.(12) gives

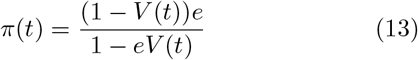

Vaccination has been reported to reduce the time until viral clearance of those infected [28]. It was reported that the time from viral peak to viral clearance was 2 days shorter for vaccinated individuals. To model this we assume that there is no viral shedding detectable by either PCR or LFD test 10 or more days after exposure, shortening the time to viral clearance considerably more than the reported effect to ensure we do not underestimate the effect of vaccines in this sensitivity analysis. For infections in vaccinated people we denote these modified functions using 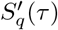, and *P*^*′*^(*τ*) for the equivalent of Eq.(6) with S’ substituted for S. The modified version of Eq.(7) is

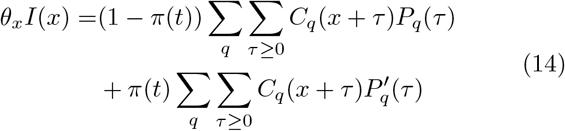

and finally the modified Eq.(8) is

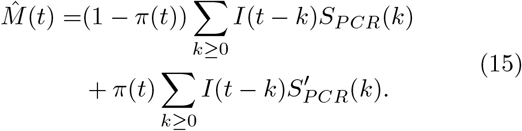

## 2 Results

The percentage of cases ascertained estimated with Eq. 8 varies by time, region, and age band (Fig. 2). These results are sensitive to variation in surveillance data, particularly when infection levels are low and there is less data to inform the estimate. For example, from the time that the infection survey began until September 2020, which is not shown in the figure, the results are highly variable and occasionally produce estimates of ascertainment that are above 100%. In general, when case rates are low we see an increase in variability due to the smaller sample size, whereas when case rates are relatively high the estimated ascertainment rate becomes more reliable.

**Figure 2:**
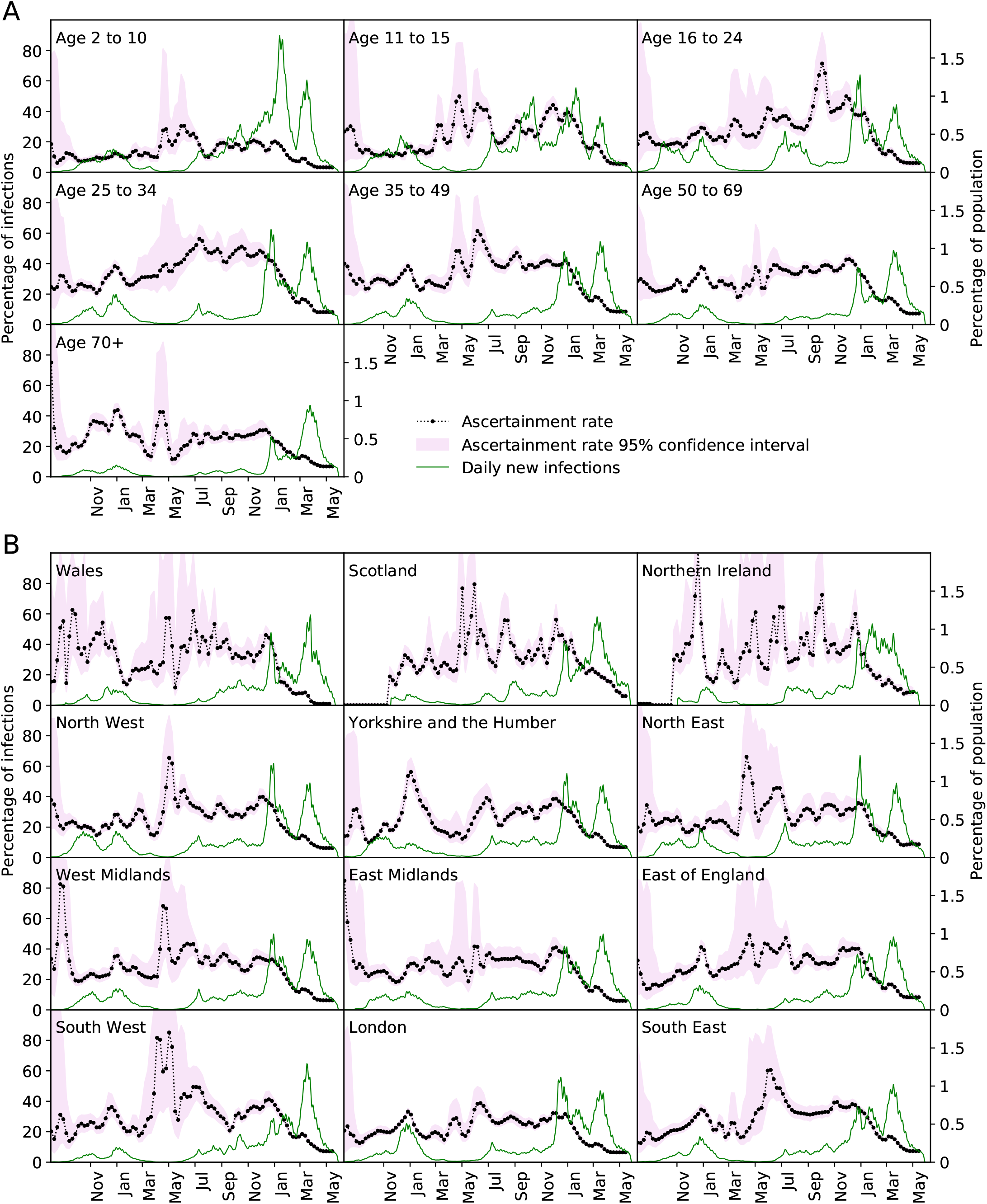
Ascertainment rate in the 7 age bands and 12 regions of the UK expressed as a percentage, from Eq. (8). Presented are the median and confidence intervals from the distribution of solutions to Eq. (8) over 200 samplings of the surveillance data, *M*(*t*). Incidence, *I*(*t*), is shown as a percentage of the population.

Case ascertainment is related to the proportion of infections that lead to symptomatic infection. This is apparent from the low ascertainment rates observed in the lowest age categories, which are known to be less likely to develop symptoms [33]. There were notable increases after March 2021 in school age children, possibly indicating that the mass testing in that age category that coincided with school reopening caused a higher detection rate of asymptomatic infections. Similarly, since vaccination is effective at preventing infections from becoming symptomatic, the decreasing ascertainment rate seen in the two highest age bands from January to April 2021 may have resulted from vaccination in those groups.

Increases from April to June 2021 occur in every group and appear to coincide with the rise in cases of the Delta variant. While this could imply that the Delta variant is more likely to cause symptomatic infection, it could also be the result of behavioural factors as restrictions to physical contact were being removed and lateral flow tests were being more widely used. There is similarity between the time series of age bands that are close to each other, whereas changes in the ascertainment rate in any given region appears to be unaffected by neighbouring regions (Fig. S3).

To compare regions, ages, and different phases of the pandemic, we consider 5 different variant classes: the wildtype that existed before the emergence of the Alpha variant, the Alpha and Delta variants, and the BA.1 and BA.2 sub-lineages of the Omicron variant, where we have used S-gene target failure to approximate the proportion of cases belonging to each class. Modelling a different time-independent ascertainment rate for each variant provides remarkable agreement between the modelled population prevalence and the value reported by the surveillance study (Fig. S1). The best-fit ascertainment rates are shown in Fig. 3. Differences between variants may reflect varying symptomatic responses, or they may reflect other behavioural factors that have changed over time.

**Figure 3:**
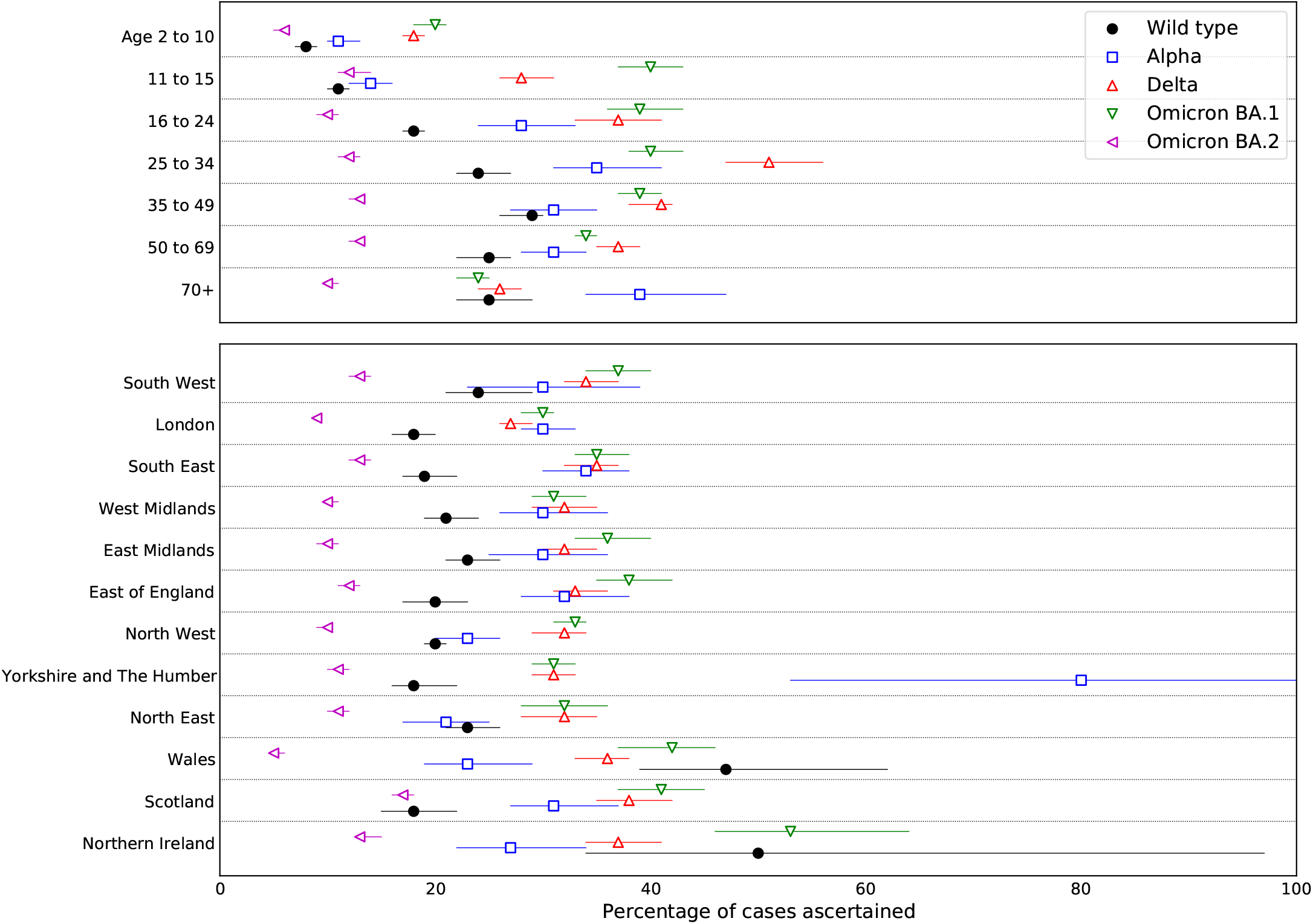
Estimates of time-independent ascertainment rates *ϕ* = (*ϕ*_*w*_, *ϕ*_*A*_, *ϕ*_Δ_, *ϕ*_*o*1_, *ϕ*_*o*2_). Presented are the median and confidence intervals from the distribution of solutions to Eq. (11) over 200 samplings of the surveillance data, *M*(*t*).

The ascertainment rate for the wild-type is lower than the rate for the Alpha, Delta and Omicron BA.1 variants across almost all age bands and regions of the UK. While the difference between Alpha, Delta and Omicron BA.1 is less clear, it is typically the case that ascertainment increased for the Delta variant over the Alpha and increased again for the Omicron BA.1 variant before decreasing substantially for the Omicron BA.2 variant during a time when free access to LFD and PCR tests was no longer available. Ascertainment rates are lowest in the youngest age band and increases with age up to the 25 to 34 band. During times when free access to testing was widely available, around 30% to 40% of cinfections were ascertained. This is lower than the percentage of infections that are symptomatic, estimated to be around 70% [34, 35], implying that a considerable number of symptomatic infections do not get ascertained.

We calculate the IFR for each month for the 4 oldest age bands (Figure 4). We have chosen not to show lower ages as the low numbers of deaths in these groups make the results highly variable do not provide a reliable estimate of the true IFR. Within the 4 age bands for which data are sufficient, the IFR increases with age. The increasing trend in IFR for the two oldest age bands in November 2020 may be a combination of higher severity of the Alpha variant [36], the increased pressure on the healthcare system, or a seasonal affect on immunity. The subsequent reduction is close to what we would expect to see given that the vaccines give some protection against infection; while vaccines reduced the number of deaths considerably, they simultaneously reduced the number of infected people. For instance, using 90% effectiveness of vaccines against death and 56% against infection [29, 37], one can calculate from the definition of effectiveness that the IFR of the vaccinated population should be 22% of the IFR for the unvaccinated.

**Figure 4:**
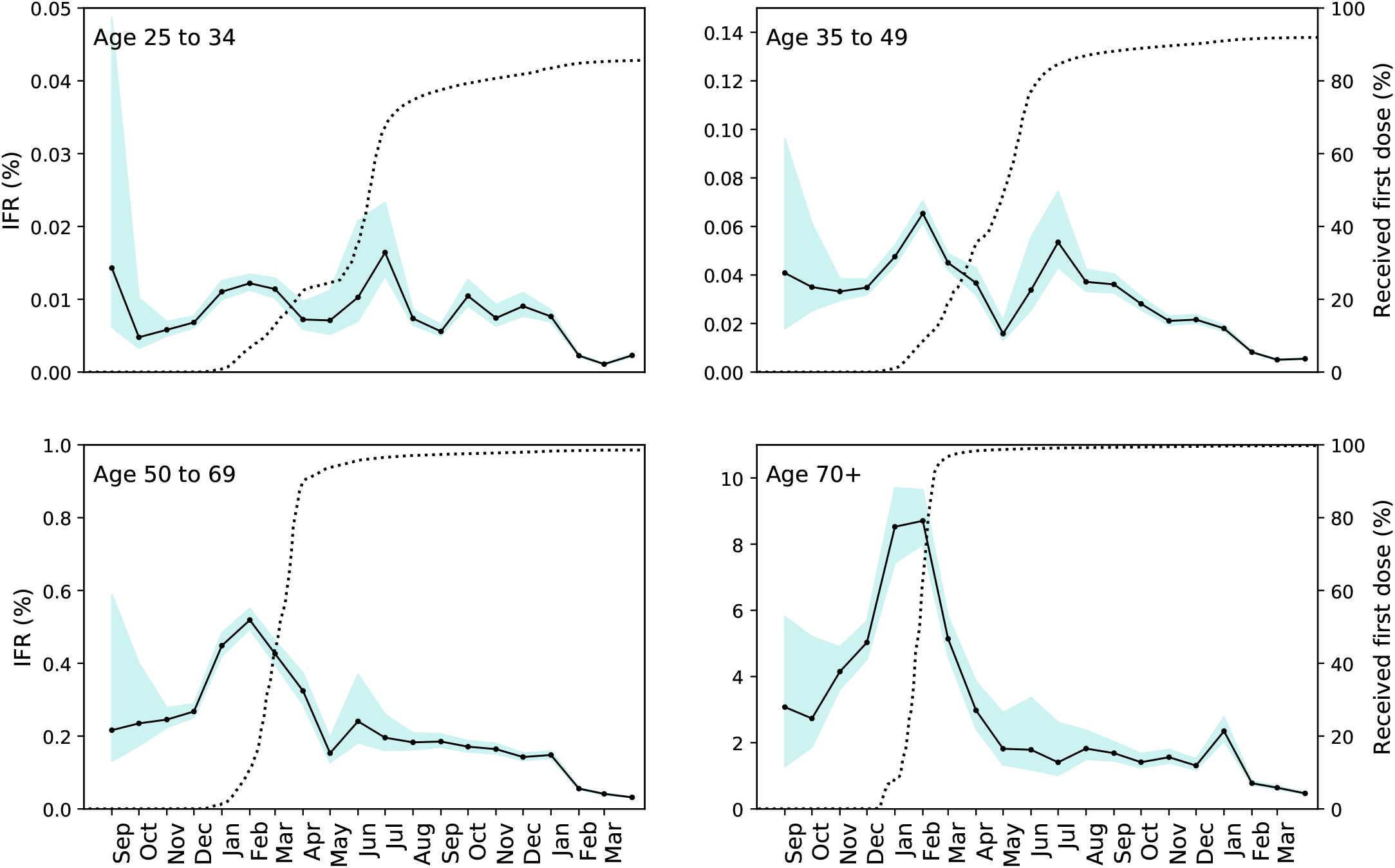
Infection fatality rate (IFR). The estimated percentage of infections that cause mortality. The shaded region shows the 95% confidence interval computed by using *I*(*x*) computed from the upper and lower estimates of prevalence given by the surveillance data. Dashed lines show the population in the respective age band that had received at least one dose of a COVID-19 vaccine.

We tested the robustness of these results against reasonable changes in the assumptions of our model. Firstly, viral clearance may occur more rapidly in individuals who have been vaccinated [28]. While we cannot model this effect precisely, making liberal assumptions about vaccine effectiveness and its effect on the test-sensitivity profile (see Section 1.1) gives results that are lower by a few percent (Fig. S2). The most substantial effect is observed in older age bands. Similarly, the IFR presented in Fig. 4 may be an overestimate during times when vaccine coverage is high. Fig. S4 shows the range of values that are plausible given the duration of viral shedding in vaccinated individuals.

Secondly, the model assumed a delay between symptom onset and receiving a PCR test of *δ*_PCR_ = 1 day. We do not have observational evidence to support this and *δ*_PCR_ = 2 is also reasonable. Repeating the analysis with *δ*_PCR_ = 2 yields a mean increase (across all age bands and variants) of 0.05 percentage points with a standard deviation of 0.89 to the results reported in Fig.3, suggesting relatively low sensitivity to this modelling decision. Thirdly, we assumed that the proportion of cases reported from LFD tests, as opposed to PCR, for England could be applied across all age bands and regions, whereas in reality they are unlikely to be proportioned equally. Repeating our analysis under the extreme assumption that 100% of community and healthcare reported cases are from LFD tests results in an mean decrease of 0.08 percentage points with a standard deviation of 1.75, again demonstrating low sensitivity to this modelling assumption.

Finally, some empirical estimates of test sensitivity are higher than the maximum of *S*_PCR_ and *S*_LFD_ [38, 39]. Repeating our analysis using an adjusted versions *S*_PCR_ and *S*_LFD_ that are linearly scaled so that *S*_PCR_ peaks at 1, we find ascertainment rates increase by a mean of 7.5 percentage points with standard deviation of 3.4, suggesting that any inaccuracy in our assumption about test sensitivities could substantially affect the outcomes presented here.

## 3 Discussion

The extensive efforts in the United Kingdom to monitor the COVID-19 epidemic have provided the opportunity to quantify a critically important parameter - the ascertainment rate - defined as the likelihood that an infected individual will get tested and receive a positive diagnosis. Here we compared the daily reported number of cases to an unbiased estimate of population prevalence to estimate the proportion of cases that are ascertained through community testing and healthcare. We also computed the daily number of new infections and from this were able to track the infection fatality ratio across time.

Variation in case ascertainment may result from differences in clinical presentation, public perception, availability of testing, or many other possible reasons. It was revealed to be related to age, with infections in the youngest age bands being the least likely to be diagnosed. Infections related to the Alpha, Delta, and Omicron BA.1 variants were more likely to be ascertained compared to variants that were circulating earlier in the pandemic (the wild type) or during a time when access to free tests was no longer available (BA.2). The IFR showed substantial variation across time, increasing substantially into winter 2020 before declining with the distribution of vaccines.

Ascertainment appears to be dependent on the SARS-CoV-2 variant. It is not possible to determine the extent to which this variation is caused by changes in symptomatic response or by external factors that may alter the propensity of the individual to seek a test. After accounting for the effects of the different variants on the ascertainment rate, we have shown that the two data sources are largely in agreement with each other. This suggests a consistency in test-seeking behaviour over time periods of months, highlighting the reliability of the diagnostic test data as measure of epidemic severity. In general, when cases are increasing, it is because infections are increasing, not because people have become more likely to receive a tests, although changes in test-seeking do occur on longer time scales.

The challenge when comparing the trend seen in random survey data to that seen in reported community cases is that the former is a measure of prevalence and the latter a measure of incidence. Our methodology resolves this by modelling the relationship between the two. Our method is related to the *deconvolution* approach previously used to estimate the incidence of other infectious diseases [40]. Indeed, this approach could be applied directly to the surveillance data to estimate incidence, however, it would not reflect changes in incidence that occur on a sub-weekly time-scale. Because our method utilises the daily resolution of the case data, it captures the daily variation in incidence while achieving almost perfect consistency between the two data sources.

The estimation of infection incidence performed here offers an alternative to methods that use serological data [41]. This allows for more accurate representation of key metrics related to epidemic control such as the reproduction rate, generation time, case doubling rates, hospitalisation and fatality rates. Our analysis revealed considerable variability in the IFR that goes beyond that expected from age and vaccination status alone. The three-fold increase in IFR in the 70+ age band beginning in November 2020 suggests that multiple factors contribute to the risk of death from infection and therefore there may be multiple ways to minimize mortality in future winter seasons. The subsequent decline adds to the body of evidence showing the effectiveness of vaccinations.

Our results are dependent on on a number of simplifying assumptions. We have applied a model that assumes all individuals experience similar viral dynamics once infected, and the time for between exposure and receiving a test follows the same distribution regardless of age or location. We have assumed that testing occurs at the time of symptom onset plus an additional delay, however, since LFD tests are expected to be used for asymptomatic screening the time between exposure and receiving an LFD test may be shorter than we have assumed. This would particularly affect children during periods when LFD testing was widely used in schools.

We highlight that the methods here may be translated to a variety of current and future epidemiological studies. As the COVID-19 pandemic has expanded the scale and scope of health surveillance data to an unprecedented level, the methods required to parse such data, and create interpretations useful to inform decision makers and increase public awareness, need also to adapt. The methods presented here are novel, although built from established mathematical concepts, and this reflects constant requirement to re-evaluate and refresh the set of mathematical and statistical tools available to analysts as the landscape of public health continues to evolve.

## Data Availability

All data are taken from publicly available sources referenced within the manuscript

https://github.com/EwanColman/Estimating_SARS-CoV-2_case_ascertainment

## Code availability

https://github.com/EwanColman/Estimating_SARS-CoV-2_case_ascertainment

---

**Figure S1:**
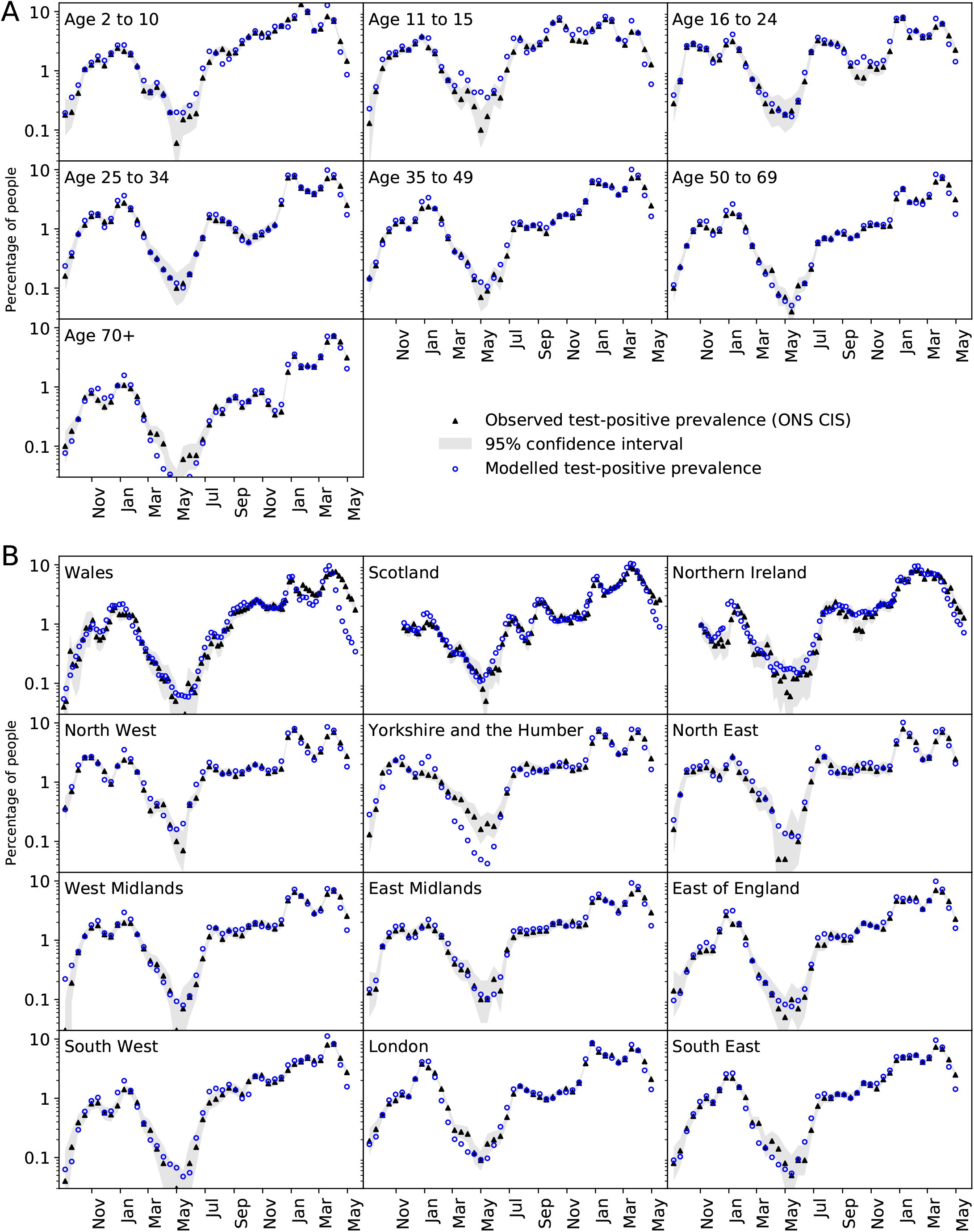
The percentage of people who would test positive if tested. Comparison of the ONS CIS to the modelled value based on reported cases and the estimated ascertainment rates given in Figure 3.

**Figure S2:**
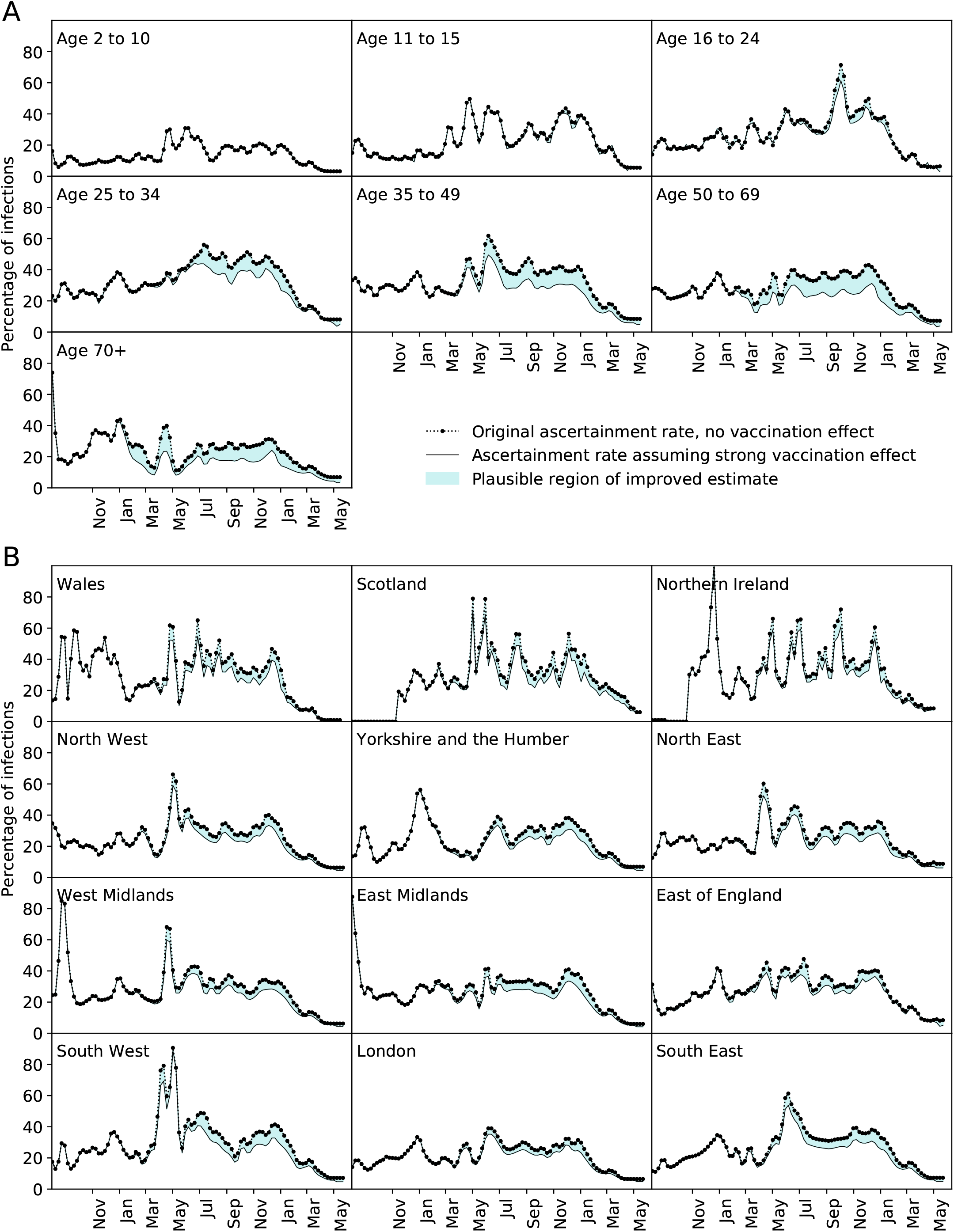
Ascertainment rates shown in Fig. 8 compared to the equivalent value that incorporates the effect of vaccination. Modelling assumptions have been made to provide the largest reasonable deviation from the original ascertainment estimate with the data available. Therefore it is likely that a precise treatment of vaccination in the model would yield a result within the shaded area between the curves.

**Figure S3:**
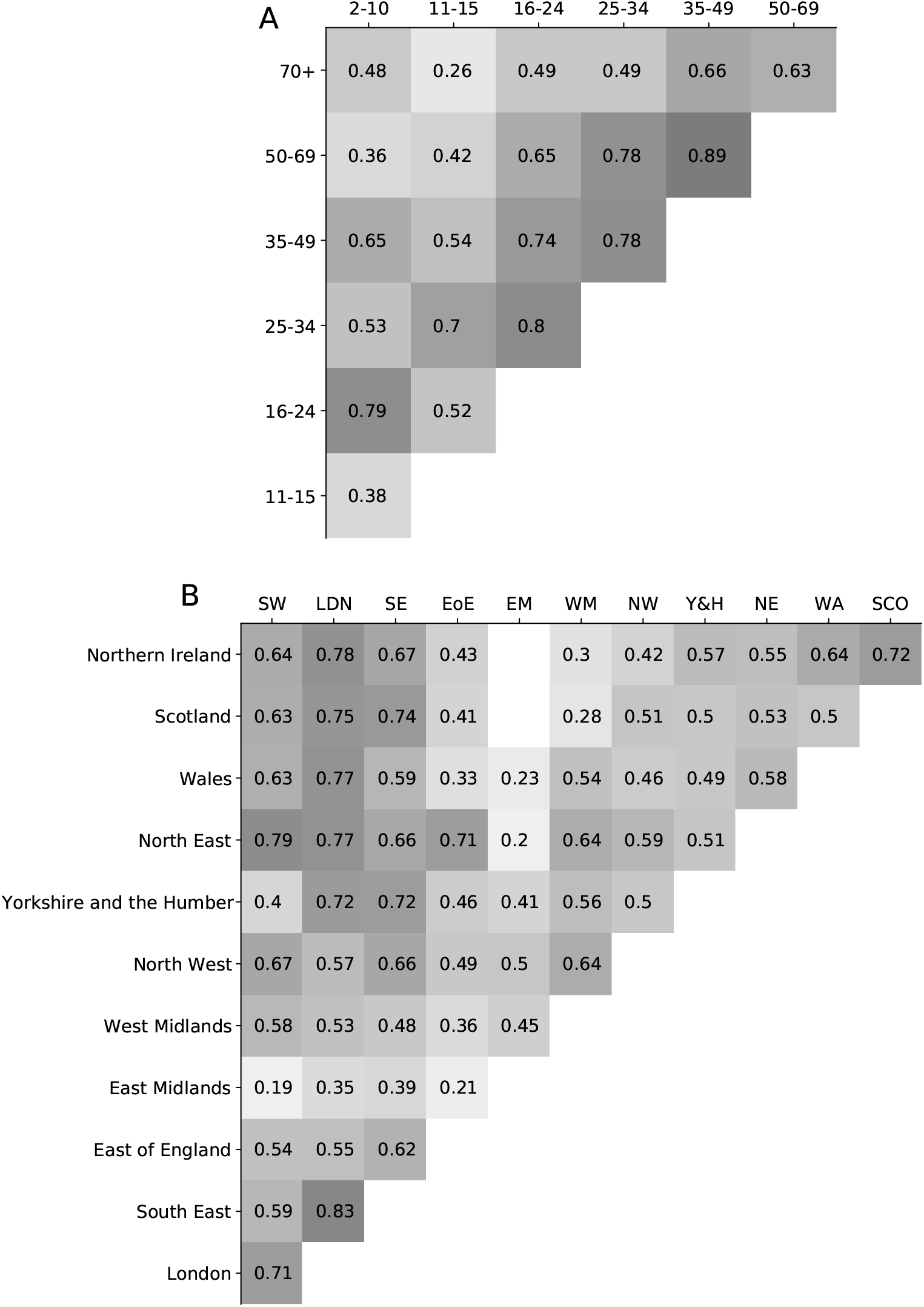
Correlations between the ascertainment rate time series of (A) all age bands and (B) all regions. Values show the Pearson correlation coefficient, *r*. Correlations where *r <* 0 or *p >* 0.05 are not displayed.

**Figure S4:**
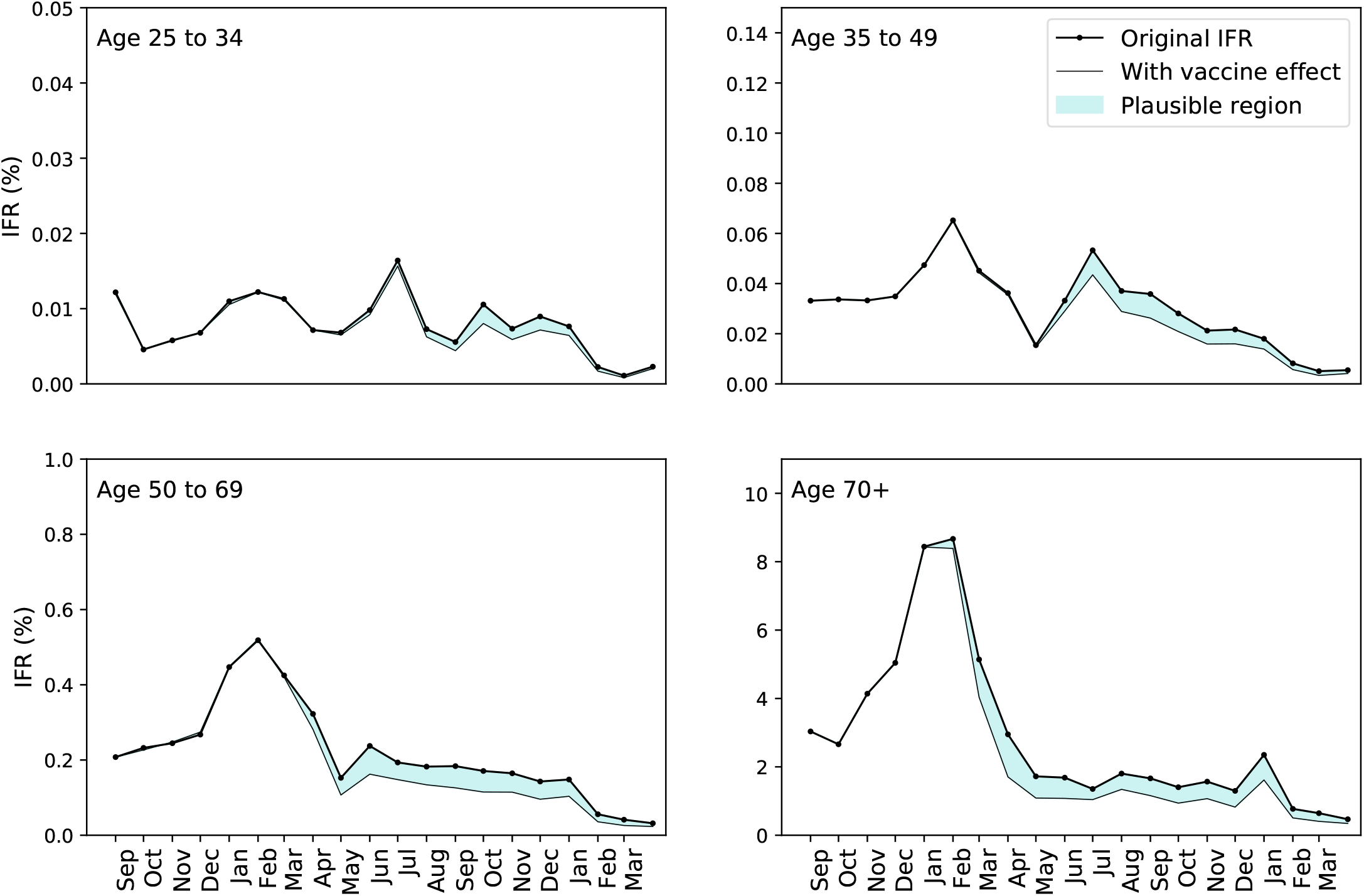
Infection fatality rates shown in Fig. 4 compared to the equivalent value that incorporates the effect of vaccination. Modelling assumptions have been made to provide the largest reasonable deviation from the original ascertainment estimate with the data available. Therefore it is likely that a precise treatment of vaccination in the model would yield a result within the shaded area between the curves.

